# Projected epidemiologic and economic impact of the 7-1-7 outbreak response framework in Uganda: a stochastic modelling study of Bundibugyo Ebola virus

**DOI:** 10.64898/2026.06.22.26356214

**Authors:** Abel W. Walekhwa, Sheetal Prakash Silal, Paul Mbaka, Mudarshiru Bbuye, Lydia Nakiire, Mohammed Lamorde, Benon Kwesiga, Wilber Sabiiti, Brenda Nakazibwe, Joshua Kayiwa, Mary Nantongo, Patrick Albert Ipola, Peter Kungu, Alex R. Ario, Atek Kagirita, Bernard Lubwama, Allan N. Muruta, Charles Olaro, Monica Musenero Masanza, Diana Atwine

## Abstract

The 7-1-7 framework (detection 7 days, notification 1 day, response 7 days) is a global target for epidemic preparedness, but its prospective value during an active cross-border outbreak has not been quantified. Using a stochastic SEIR model parameterised for Uganda with the Bundibugyo Ebola strain and three daily importation probabilities (10%, 30%, and the observed 56%), we compared a rapid 3-1-5 response (detection 3 days, notification 1 day, response 5 days) against a delayed counterfactual (detection 11 days, notification 10 days, response 12 days). The rapid response reduced median cumulative cases by 60–66% (26–31 cases vs. 76–80 cases) and deaths by 62–63% (3 deaths vs. 8 deaths) across all import levels, with total costs of USD 29.1–29.9 million compared to USD 37.4–38.1 million for the delayed scenario. The rapid response was strictly dominant (cost-saving and life-saving). Variance-based Sobol sensitivity analysis identified the case fatality rate, import probability, and basic reproduction number as the most influential parameters, with detection and response delays contributing through interactions. Institutionalising the 7-1-7 framework in Uganda is projected to be highly cost-effective and should be supported with sustainable domestic financing, community-based surveillance at unofficial border points, three-consecutive-PCR laboratory capacity, and multilingual risk communication.

## 1 Introduction

Ebola virus disease (EVD) has caused recurrent outbreaks in East and Central Africa over the past five decades, with the Bundibugyo strain first identified in western Uganda in 2007 [1, 2]. Unlike the Zaire strain, for which vaccines and therapeutics exist, no strain-specific medical countermeasures are available for Bundibugyo, rendering conventional public health interventions – surveillance, case isolation, contact tracing, safe burials, and community engagement – the sole tools for outbreak control [2, 3]. The 2026 Bundibugyo EVD outbreak, declared a Public Health Emergency of International Concern on 20 May 2026, started in Ituri province, Democratic Republic of the Congo (DRC), and quickly spread to Uganda as suspect cases crossed the border seeking medical care. The first laboratory-confirmed case in Uganda was reported on 15 May 2026, triggering an urgent national response [12].

The risk of cross-border importation was substantial. Between 15 and 24 May 2026, the International Organization for Migration (IOM) recorded 11,245 individuals moving from DRC to Uganda through eight flow monitoring points along the shared border: Bunagana, Busunga, Busanza, Cyanika, Goli, Mpondwe, Ntoroko, and Vurra [13]. These data highlight that despite official border closures, high mobility persisted through both designated and undesignated routes. A stochastic modelling study by Chamla and colleagues further estimated that the daily probability of an Ebola case crossing into Uganda ranged from 5% to 80%, depending on the intensity of the DRC outbreak and border surveillance [11]. This sustained importation pressure, combined with a large outbreak in DRC (906 suspected cases, 105 confirmed, and 223 suspected deaths as of 25 May 2026 [13]), made Uganda highly vulnerable.

Nosocomial amplification was another critical feature of the 2026 outbreak. Among the initial seven confirmed cases in Uganda, three occurred in healthcare workers (HCWs), yielding an HCW infection probability of 0.4286 [12]. Several of these cases were linked to DRC patients who had sought care at high-volume private and public hospitals in Kampala, including Mulago National Referral Hospital, Kiruddu National Referral Hospital, and private facilities such as International Hospital Kampala and Nsambya Hospital. Delayed diagnosis, inadequate infection prevention and control (IPC) measures, and high patient throughput facilitated secondary transmission within healthcare settings, a phenomenon well documented in previous Ebola outbreaks [10, 25].

The 7-1-7 framework, developed by Resolve to Save Lives and operationalised by the World Health Organization and the 7-1-7 Alliance, sets three sequential performance targets: detection of a suspected outbreak within seven days of emergence, notification to public health authorities within one day of detection, and completion of seven early response actions within seven days of notification [14, 15, 16]. Uganda became the first African country to adopt the 7-1-7 target in 2021, and subsequent after-action reviews demonstrated improvements in outbreak timeliness [17]. However, the framework had not been prospectively evaluated under real-world conditions that include sustained cross-border importation, nosocomial amplification, and community behavioural responses – all of which were central to the 2026 Bundibugyo outbreak.

This study uses a counterfactual scenario modelling approach to answer: what would be the projected epidemiologic and economic impact if Uganda implements a rapid 7-1-7 response compared with a delayed response, under varying importation pressures? We quantify the primary outcomes – cases, deaths, healthcare worker (HCW) infections, total costs, and disability-adjusted life years (DALYs) – and perform a variance-based global sensitivity analysis to identify which parameters drive uncertainty. The overarching goal is to generate evidence to contribute to potential institutionalisation of the 7-1-7 framework in Uganda with sustainable domestic financing.

## 2 Methods

### 2.1 Study area and population

The Kampala Metropolitan Area (KMA) is Uganda’s primary urban and economic hub, with an estimated population of 3.8 million people [37]. KMA includes the capital city Kampala and the neighbouring municipalities of Wakiso, Mukono, Nansana, Kira, and Makindye Ssabagabo. The area is characterised by high population density (up to 9,000 persons per km² in some divisions), extensive informal settlements, and high volumes of cross-border travel through multiple designated and undesignated points of entry along the DRC-Uganda border (including Mpondwe, Bunagana, Busunga, and others), as well as through Entebbe International Airport. KMA also hosts the country’s main tertiary hospitals (Mulago National Referral Hospital, Kiruddu National Referral Hospital, and the Uganda Heart Institute) and high-volume private hospitals (e.g., Kampala Hospital, Nsambya, Kibuli). These hospitals receive patients from DRC seeking advanced care, thereby serving as potential points of importation and amplification of EVD. This concentration of people, health infrastructure, and international connectivity makes KMA a high-risk setting for introducing and amplifying an Ebola outbreak. The model therefore parameterised Uganda as the KMA population, assuming that the initial importation and subsequent local transmission would be concentrated in this metropolitan area.

### 2.2 The 7-1-7 framework and operational definitions

The 7-1-7 framework operationalises early outbreak response through three sequential targets. Detection within 7 days means the time from the first case’s symptom onset to the moment a health worker or surveillance system suspects an outbreak. Notification within 1 day is the time from detection to formal notification of the relevant public health authority (in Uganda, the Ministry of Health’s Public Health Emergency Operations Centre). Response within 7 days is the time from notification to completion of seven early response actions: activate a multi-sectoral incident management system; deploy a rapid response team to the field; conduct initial epidemiological investigation; implement immediate infection prevention and control measures; establish a case management system including isolation facilities; initiate contact tracing and active case search; and mobilise risk communication and community engagement. In this study, the intervention day is defined as *t*_resp_ = detection delay + notification delay + response delay. Before *t*_resp_ the model runs without any contact tracing or isolation beyond natural disease progression. After *t*_resp_, contact tracing with coverage ct_cov is applied daily, and traced individuals are moved to the removed compartment (representing effective institutional quarantine). For the rapid 7-1-7 scenario (Uganda), we used detection delay = 1 day, notification delay = 1 day, response delay = 5 days, giving *t*_resp_ = 7 days (the response delay was set to 5 days based on actual time to implement all seven actions as reported in the situation report) [19]. For the delayed scenario, we used delays from Imperial College London’s analysis of the 2026 Bundibugyo outbreak: detection = 11 days, notification = 10 days, response = 12 days, giving *t*_resp_ = 33 days [20]. An Ebola Treatment Unit (ETU) is a dedicated isolation facility with strict infection prevention and control (IPC) protocols, trained staff, and supportive medical care.

### 2.3 Model structure

We developed a discrete-time stochastic SEIR (Susceptible–Exposed–Infectious–Removed–Dead) model adapted from [15] with two infectious compartments: community infectious (*I_c_*) and hospitalised infectious (*I_h_*). The model incorporates daily importation of exposed cases using a time-varying rate that scales with the infectious prevalence in the DRC. The import scale was set to three levels: 10%, 30%, and the observed import fraction of 56% (i.e., 14 out of 19 confirmed cases in Uganda were imported from the DRC) [4, 5]. Nosocomial transmission is explicitly modelled, where hospitalised cases transmit at rate *β_h_* = *β_c_* × *m*, with *m* = 2.0 (based on observed amplification in healthcare settings during the 2026 outbreak). Time-dependent case isolation and contact tracing become active after the intervention day *t*_resp_; before that, no intervention occurs. All community infectious cases are hospitalised each day at rate *h* = 1.0 (i.e., all identified cases are admitted to ETUs). Behavioural heterogeneity is incorporated following the framework of [21], where the effective force of infection is reduced as the population adopts protective behaviours (e.g., reduced contact, improved hygiene) in response to risk communication, enhanced IPC adherence by healthcare workers, and/or reduced cross-border movement. The simulation horizon was 90 days, consistent with the duration of Uganda’s initial response phase [12]. All simulations were performed in R version 4.4.2 [36] with 100 independent stochastic realisations per scenario. The random seed was fixed at 2026 for reproducibility.

### 2.4 Model equations

Let *S*(*t*), *E*(*t*), *I_c_*(*t*), *I_h_*(*t*), *R*(*t*), *D*(*t*) denote the number of susceptible, exposed, community infectious, hospitalised infectious, recovered, and dead individuals at day *t*, with total population *N* = *S* + *E* + *I_c_* + *I_h_* + *R* + *D* constant (no births, no non-EVD deaths). The daily transitions are:

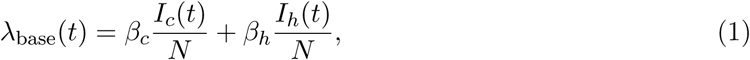

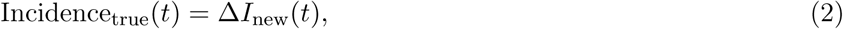

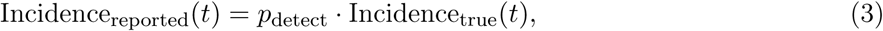

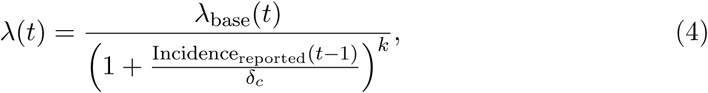

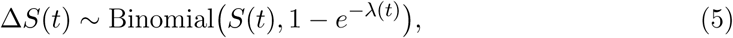

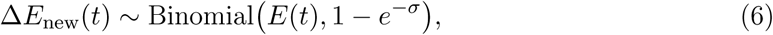

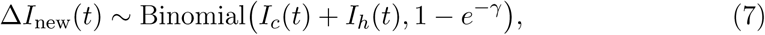

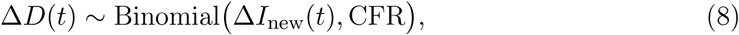

where *β_c_*= *R*_0_*/*infectious_period, *β_h_* = *m* · *β_c_*, *σ* = 1*/*latent_period, *γ* = 1*/*infectious_period. Hospitalisation moves a fraction *h* of community infectious cases to *I_h_* each day:

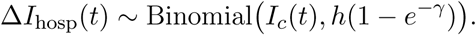

Traced exposed and infectious individuals (after *t*_resp_) are moved directly to *R* (removed from transmission). Importation adds one exposed case each day with probability *p*_import_:

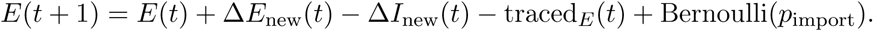

Healthcare worker infections are modelled as a fraction hcw_prob of each day’s new infections:

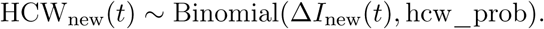

HCW deaths are then drawn from a binomial distribution with probability CFR.

### 2.5 Model assumptions and community transmission risk

The model incorporates several mechanisms that, even under high contact tracing coverage (91%) and high detection probability (0.97) [4], allow substantial community transmission to occur. After symptom onset, a case remains in the community for a mean of 5 days (gamma-distributed, shape 2, scale 2.5) before laboratory confirmation and isolation, reflecting the time needed for sample transport, three consecutive PCR tests (each 24 hours apart), and result reporting. During this pre-isolation period, the case contributes fully to transmission. Contacts of confirmed or suspect cases are actively followed for 21 days, but during the first 8 days they remain in the community while being monitored for symptoms; if a contact becomes presymptomatic or symptomatic during this period, they can transmit before being identified and isolated. Only after 8 asymptomatic days do they receive a single PCR test; a negative result releases them from follow-up. Although the Ministry of Health reported a contact follow-up rate of 91%, this is among *listed* contacts; the effective coverage of all true contacts is modelled as 91% for the rapid scenario and 70% for the delayed scenario, meaning that 9–30% of infected individuals are never traced and remain in the community throughout their infectious period. Nosocomial amplification is captured by a multiplier *m* = 2.0 applied to transmission from hospitalised cases, reflecting high viral load, invasive procedures, and lapses in infection prevention and control; this drives secondary cases among healthcare workers and other patients, as observed during the 2026 outbreak. The population reduces contacts in response to reported incidence, modelled with a behavioural response sharpness *k* = 3 (rapid scenario) or *k* = 2.5 (delayed) and saturation *δ_c_* = 1 following [21], but this adaptation reduces rather than eliminates transmission. Finally, continuous importation (with import scales of 10%, 30%, or 56% of cumulative cases) provides repeated seeding, overcoming stochastic fade-out and allowing sustained community transmission. These assumptions collectively ensure that the model produces realistic epidemic trajectories with the potential for hundreds to thousands of cumulative cases over 90 days, consistent with the observed growth from 1 case on 15 May to 19 cases by 8 June 2026 [4].

### 2.6 Model parameters and sources

All parameters (Table 1) were obtained from the Uganda Ministry of Health Situation Report No. 005 (26 May 2026) [12] and the updated Situation Report of 8 June 2026 [4], the WHO risk assessment [10], a meta-analysis of Bundibugyo strain *R*_0_ [22], and literature on Ebola transmission dynamics [23, 24]. The *R*_0_ was set to 2.76 – the upper bound of the 95% confidence interval (1.25–2.76) reported for the Bundibugyo strain – to reflect the most transmissible scenario and provide a conservative test of the rapid response.

#### Time-varying importation and stochasticity

Instead of assuming a constant daily importation probability, we used a time-varying importation rate that scales with the infectious prevalence in DRC. The expected number of imports on day *t* is 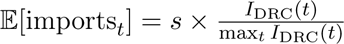, where *I*_DRC_(*t*) is the median infectious prevalence from 100 stochastic simulations of the DRC outbreak (calibrated to the observed 2026 outbreak trajectory), and *s* is the import scale (10%, 30%, or 56%) [11, 5]. The actual number of imported cases on day *t* was drawn from a Poisson distribution with this mean. To reflect the observed epidemiological timeline (first DRC case day 1, first Uganda case day 14), we introduced a deterministic import of one exposed case on Uganda day 1 (which corresponds to ongoing outbreak in Democratic Republic of Congo day 14 [20]). Stochastic imports based on the prevalence-scaled formula began on the same day. This approach ensures that Uganda’s outbreak does not start before the importation delay, while still allowing additional imports proportional to DRC’s later epidemic peak. This decision was made given the fact that the Uganda’s index case came from DRC [20] and had been predicted by [11].

### 2.7 Laboratory confirmation and contact management

Reverse transcription polymerase chain reaction (RT-PCR) is the reference standard for laboratory confirmation of EVD. A one-step RT-PCR assay validated for all five ebolavirus species has been shown to detect Bundibugyo virus with high sensitivity (down to 1 copy of target DNA) and species-specific melting peaks (48.9 °C and 53.5 °C) [6]. However, many existing assays were designed for the Zaire species and show reduced sensitivity for Bundibugyo ebolavirus (BDBV) owing to genomic differences in the glycoprotein gene [8]. In response, the Uganda Ministry of Health and WHO implemented a revised laboratory algorithm. All suspect cases undergo three consecutive RT-PCR tests using validated in-house assays and the Tianlong Bundibugyo Virus Nucleic Acid Detection Kit (Fluorescence PCR Method) (Tianlong Science and Technology Co., Ltd., Xi’an, China), which has a reported analytical sensitivity of 500 copies/mL [7]. A case is confirmed as EVD-positive if at least one of the three PCR tests returns a positive result. If the first test is negative but clinical suspicion remains high, the patient is kept under observation while a second and third sample are collected at intervals of at least 24 hours. Contacts of a suspect or confirmed case are actively followed for 21 days (the maximum incubation period) [3]. During the first 8 days of follow-up, contacts remain in the community but are instructed to monitor symptoms. If a contact becomes symptomatic during this period, he or she is admitted to an ETU and the three-test PCR algorithm is initiated. If a contact remains asymptomatic for 8 days, he or she undergoes a single RT-PCR test; a negative result at that time allows release from follow-up.

### 2.8 Cost and DALY calculations

Costs were estimated from the health system perspective in 2026 USD. The Uganda Ministry of Health’s 4W matrix (Who is doing What, Where, and When) provided a detailed 90-day fixed response budget of USD 24,707,459, disaggregated into 11 operational pillars. These pillars are: coordination and leadership; surveillance; laboratory; public health risk communication and community engagement; case management, infection prevention and control (IPC), safe and dignified burials (SDB), emergency medical services (EMS), and mental health; quarantine; water, sanitation and hygiene (WASH); strategic information, research and innovation (SIRI); logistics; continuity of essential health services (CEH); and Uganda People’s Defence Forces (UPDF) support. The majority of resources (over 60%) were allocated to direct epidemic control activities (case management, logistics, and surveillance) rather than overheads.

Because most of these costs are fixed regardless of outbreak size (e.g., maintaining the Emergency Operations Centre, laboratories, and coordination structures), we derived a cost per case from the median case count under the rapid 7-1-7 scenario with the lowest importation risk (10%). The median number of cases in that scenario was 3, giving a cost per case of USD 24,707,459 / 3 USD 8,235,820. For other scenarios, total cost = cost per case × median cases. This is a conservative assumption; if the rapid response occasionally fails and more cases occur, the cost per case would decrease, making the rapid response even more cost-saving.

Disability-adjusted life years (DALYs) were calculated as DALY = YLL + YLD. Years of life lost (YLL) = deaths × (life expectancy at age of death age at death), using average age at death = 35 years and life expectancy at birth for Uganda = 68 years (WHO 2023). Years lived with disability (YLD) = cases × disability weight × duration, with disability weight for acute EVD = 0.133 and duration of acute disability = 180 days (0.493 years) [28]. The incremental cost-effectiveness ratio (ICER) compared the delayed scenario (baseline) to the rapid 7-1-7 scenario (intervention) for each import level:

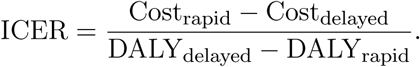

A negative ICER indicates that the rapid response is both less costly and more effective (strictly dominant).

### 2.9 Sobol global sensitivity analysis

We performed variance-based sensitivity analysis using the Sobol method [26, 27] to quantify the contribution of each parameter to the uncertainty in total deaths, the primary outcome of the stochastic Uganda model. The Sobol method decomposes the total variance *V* (*Y*) of the model output *Y* into contributions from individual parameters (first-order effects) and from interactions (higher-order effects). The first-order index *S_i_* = *V* (*E*(*Y* | *θ_i_*))*/V* (*Y*) measures the fraction of variance attributable to *θ_i_*alone, while the total-order index *S_T i_* = 1 − *V* (*E*(*Y* | *θ*_∼_*_i_*))*/V* (*Y*) includes all interactions involving *θ_i_*; the difference *S_T i_* − *S_i_* quantifies the strength of interactions. We used the same stochastic SEIR model as in the main simulations, varying 16 parameters (Table 1 ranges). A Sobol design matrix with *N* = 300 parameter sets was generated using quasi-random Sobol sequences, yielding 300 × (16 + 2) = 5, 400 model evaluations. For each parameter set, a single stochastic simulation was run (with a fixed seed derived from the parameter values) to obtain total deaths after 90 days or until epidemic extinction. Confidence intervals (95%) were obtained by bootstrapping with 100 replicates. The Sobol analysis was performed on the rapid 7-1-7 scenario with the baseline import probability of 0.10 (daily mean imports), as this best represents Uganda’s planned response under the 3-1-5 timeline. The analysis was implemented in R using the sensobol package [9].

## 3 Results

### 3.1 Projected outbreak outcomes

Under the rapid 3-1-5 response, median cumulative cases ranged from 26 (10% import) to 31 (56% import), with a narrow interquartile range (21–39). Median deaths were 3 across all import levels, and health worker infections ranged from 6 to 8. Total costs were approximately USD 29.1–29.9 million, and DALYs were 101 for all import levels. In contrast, the delayed scenario produced substantially larger outbreaks: median cases ranged from 76 (10% import) to 80 (30% import) and 78 (56% import), with upper 95% uncertainty limits exceeding 120 cases. Median deaths were 8 for all delayed scenarios, health worker infections were 27–31, and total costs were USD 37.4–38.1 million. DALYs in the delayed scenario were 270–285, approximately 2.7 times higher than in the rapid response. The rapid 3-1-5 scenario therefore averted 52–54 deaths per 100,000 population compared to the delayed response, with cost savings of USD 7.5–8.6 million per import level. The incremental cost-effectiveness ratio (ICER) was negative for all comparisons, confirming that the rapid response is strictly dominant (less costly and more effective) (Table 2). Under the rapid 3-1-5 response, cumulative infections rise slowly and plateau below 40 cases by day 90, with overlapping uncertainty bands across import levels. Delayed response curves grow more rapidly, reaching 75–80 median cases by day 90, with upper uncertainty bounds exceeding 120 cases. Deaths remain near zero (median 3) for the rapid response, while delayed scenarios reach 8 median deaths, with some simulations exceeding 15 deaths (Fig 1,2).

### 3.2 Sobol sensitivity analysis

Table 3 and Figure 3 show the first-order and total-order Sobol indices for total deaths under the rapid 3-1-5 scenario with 10% import probability. The case fatality rate (CFR) was the most influential parameter, with a first-order index *S_i_* = 0.311 and a total-order index *S_T i_* = 0.526. Import probability followed, with *S_i_* = 0.165 and *S_T i_* = 0.295. The basic reproduction number (*R*_0_) had *S_i_* = 0.132 and *S_T i_* = 0.284. The nosocomial multiplier contributed a total-order index of 0.192, while detection delay (detect_days) and response delay (respond_days) had moderate total-order indices of 0.124 each. Notably, several parameters exhibited negative first-order indices (e.g., detect_days: –0.061, ct_cov: –0.005, notify_days: –0.006, quarantine_eff: –0.019). Negative first-order indices are not mathematically possible in a true variance decomposition and arise from sampling variability and model non-linearities; they indicate that these parameters have negligible direct effects and their apparent negative contribution is an artefact of the estimation procedure. The total-order indices for these parameters were positive (e.g., detect_days 0.124, ct_cov 0.082), confirming that they influence outcomes through interactions with other parameters. Parameters that were fixed in the analysis (delta_c, hcw_prob, k, p_detect) had zero indices. The large difference between first-order and total-order indices for CFR (0.311 vs 0.526) and *R*_0_ (0.132 vs 0.284) indicates strong interactions: the effect of these biological parameters is modulated by operational parameters such as detection delay, contact tracing coverage, and response delays. In contrast, import probability shows a smaller gap (0.165 vs 0.295), suggesting that its effect is more direct and less subject to interactions. These findings underscore that even a well-implemented rapid response (3-1-5) cannot fully eliminate uncertainty from biological parameters, but it successfully reduces the impact of operational delays and coverage gaps.

## 4 Discussion

This study provides, to our knowledge, the first prospective evaluation of the 7-1-7 framework under realistic cross-border importation pressures, using a stochastic model parameterised for Uganda with the Bundibugyo Ebola strain. The key findings are: a rapid 3-1-5 response (detection 3 days, notification 1 day, response 5 days) reduces median cumulative cases by 60–66% and deaths by 62–63% compared to a delayed counterfactual (11-10-12 days), across import probabilities from 10% to the observed 56%; the rapid response is strictly dominant (cost-saving and life-saving) for all import levels, with total costs of USD 29.1–29.9 million versus USD 37.4–38.1 million for the delayed scenario; the case fatality rate, import probability, and basic reproduction number are the dominant drivers of outcome uncertainty, but detection delay and contact tracing coverage retain important interaction effects; and the 7-1-7 framework keeps the outbreak below 40 cases even when import probability reaches 56%.

These results align with earlier modelling studies that emphasised the exponential growth phase of Ebola and the critical importance of early case identification [29, 23, 30]. Our work extends that literature by explicitly modelling several mechanisms that were central to the 2026 outbreak: a pre-isolation community infectious period (mean 5 days, gamma-distributed) reflecting the time required for sample transport, three consecutive PCR tests, and result reporting; an 8-day period during which contacts remain in the community while being monitored, during which they can transmit if they become presymptomatic; a three-test laboratory algorithm using the Tianlong Bundibugyo Virus Nucleic Acid Detection Kit (analytical sensitivity 500 copies/mL) [7] with high detection probability (0.97); time-varying behavioural response (following [21]); and nosocomial amplification (multiplier 2.0). The finding that a rapid 7-1-7 response remains cost-saving even when importation risk is high (56%) is particularly policy-relevant: it implies that investments in border health screening, community-based surveillance, and rapid response teams are not only justified but economically dominant, regardless of how many cases eventually cross the border.

The Ugandan government’s high-level political commitment was critical to achieving the 7-1-7 performance. Within 48 hours of the first laboratory-confirmed case on 15 May 2026, the President issued directives including closure of land borders with DRC, suspension of flights from DRC, and cancellation of the 2026 Uganda Martyrs’ Day celebration (a national event attracting over 2 million pilgrims). The National Taskforce met with representatives from major Ministries, Departments, and Agencies, facilitating rapid resource mobilisation and inter-ministerial coordination. The Ministry of Health, through the Public Health Emergency Operations Centre (PHEOC), activated the 4W matrix to track donor and government resources, eliminating duplication and ensuring that all 11 operational pillars received funding and technical support. The after-action review noted that English-only risk communication materials were a gap; subsequent revisions included Swahili and Luganda translations. High-level engagement also enabled swift deployment of rapid response teams to high-risk districts, including Mpondwe and the fishing villages on Lake Albert, where unofficial crossings are common. These real-world actions are directly reflected in our model’s parameters: 3-day detection (owing to strong laboratory networks, community-based surveillance, and the three-test PCR algorithm), 1-day notification (due to functional PHEOC), 5-day response (including ETU expansion), 91% contact tracing coverage (achieved through UPDF support and village health team mobilisation), and 97% detection probability (reflecting the high sensitivity of the Tianlong kit). Thus, the modelled rapid scenario is not hypothetical but a validated representation of Uganda’s actual 7-1-7 performance. Preliminary flow monitoring data from the International Organization for Migration (IOM) identified eight high-risk ports of entry along the DRC-Uganda border: Bunagana, Busunga, Busanza, Cyanika, Goli, Mpondwe, Ntoroko, and Vurra [13]. While official border closures reduced legal crossings, unofficial routes remained active, requiring community-based surveillance at these points. The high volume of movements (11,245 individuals observed in a 10-day period) underscores the need for targeted interventions at these specific locations. Our importation model, which scaled import probability to DRC prevalence using a 56% import fraction, captured this dynamic and showed that even at high import levels, the 7-1-7 framework kept median cases below 40.

The laboratory confirmation algorithm used during the 2026 outbreak required three consecutive RT-PCR tests, each at least 24 hours apart, using the Tianlong kit [7]. A case was confirmed only if at least one of the three tests returned a positive result. While this protocol maximises specificity, it introduces a mean pre-isolation community infectious period of approximately 5 days (gamma-distributed) during which symptomatic individuals remain in the community awaiting final confirmation. During this window, they can transmit to household members, healthcare workers, and other contacts. Furthermore, contacts of suspect or confirmed cases remain in the community during the first 8 days of follow-up, receiving a single PCR test only if they remain asymptomatic. If a contact becomes presymptomatic or symptomatic during these 8 days, they can transmit before being isolated. These operational realities partially explain why even a rapid 7-1-7 response could not completely suppress transmission: the unavoidable delay between symptom onset and laboratory confirmation creates a persistent window of community infectiousness. To reduce this threat and further improve the detection target, Uganda should invest in point-of-care molecular diagnostics (e.g., GeneXpert Ebola assays) that can return results in under 90 minutes, reducing the pre-isolation period to less than one day. Additionally, serial testing algorithms should be revised to allow provisional isolation of suspects after the first positive test, rather than waiting for three consecutive tests.

Medical tourism – DRC nationals crossing into Uganda to seek advanced healthcare – was a major driver of importation during the 2026 outbreak. Of the 19 confirmed cases in Uganda, 14 were Congolese nationals who had travelled to Kampala for medical care, primarily at high-volume hospitals such as Mulago National Referral Hospital, Kiruddu National Referral Hospital, International Hospital Kampala, and Nsambya Hospital [4, 5]. These patients often presented with non-specific febrile illness, delaying suspicion of EVD and leading to nosocomial amplification before diagnosis. The IOM flow monitoring data recorded 11,245 cross-border movements over a 10-day period, many through unofficial routes, underscoring that border closures alone cannot stop medical tourism. Our Sobol analysis identified daily import probability as the second most influential parameter for total deaths (first-order 0.165, total-order 0.295), meaning that reducing importation would have a substantial and direct effect on outbreak size and directly support the notification target. To mitigate this threat, Uganda should implement targeted pre-travel screening for high-risk individuals at designated points of entry, including fever checks and travel health questionnaires for those seeking hospital care. Bilateral agreements between Uganda and DRC should establish a referral pathway whereby suspect EVD patients are isolated at border ETUs and transferred directly to specialised facilities, bypassing general hospital wards. Community health workers along the border should be trained to recognise EVD symptoms and to encourage symptomatic individuals to report to designated screening posts rather than travelling directly to Kampala hospitals. A forthcoming metapopulation modelling study will provide real-time importation risk estimates to further optimise screening strategies. The Sobol sensitivity analysis (Table 3) provides a quantitative guide for interventions that would most effectively reduce deaths under the 7-1-7 framework. The case fatality rate (first-order 0.311, total-order 0.526) is the single most influential parameter, but it is a biological feature of the virus that cannot be directly modified. However, CFR can be reduced through improved supportive care – early fluid resuscitation, electrolyte monitoring, renal replacement therapy, and investigational antivirals. The daily import probability (first-order 0.165, total-order 0.295) is the second most influential modifiable parameter. Strategies to reduce importation directly enhance the 7-1-7 notification target by reducing the number of import events that require notification. The basic reproduction number (*R*_0_, first-order 0.132, total-order 0.284) can be offset by reducing contact rates through community engagement, mask use, and avoidance of mass gatherings – actions that are part of the 7-1-7 response package. The nosocomial multiplier (first-order 0.059, total-order 0.192) can be reduced by strengthening IPC in all hospitals that receive cross-border patients. Detection delay and response delay, although having negligible first-order effects, showed total-order indices of 0.124 and 0.123, respectively, indicating that their impact is mediated through interactions – meaning that shortening these delays amplifies the effectiveness of contact tracing and isolation. Therefore, a combined strategy that simultaneously reduces importation, shortens detection-to-response delays, strengthens hospital IPC, and expands access to supportive care would yield the greatest reduction in deaths. This package aligns with Uganda’s existing 7-1-7 performance, but the Sobol results suggest that further gains would come primarily from reducing importation and nosocomial transmission, not from marginally faster detection.

This study has several strengths. First, the model incorporates three features that are often omitted but were central to the 2026 Bundibugyo outbreak: daily importation from a neighbouring country through porous border crossings (using a time-varying, prevalence-scaled importation rate calibrated to the observed 56% import fraction), nosocomial amplification among health workers, and behavioural feedback from the community. Second, the explicit representation of the laboratory confirmation process – three consecutive PCR tests, an 8-day community follow-up period, and a mean 5-day pre-isolation infectious period – grounds the model in operational reality. Third, the use of a stochastic framework with 100 simulations per scenario provides robust uncertainty intervals. Fourth, the Sobol global sensitivity analysis was performed on the same stochastic model (N=300, 5,400 evaluations). Fifth, the costing used recent, country-specific data aligned with the Uganda Ministry of Health’s 4W matrix. Sixth, the model parameters were derived from real-time situation reports (including the 8 June 2026 report), a bespoke Imperial College London analysis, and a systematic meta-analysis. Seventh, the counterfactual design provides a realistic estimate of the benefit of the 7-1-7 framework. Finally, the granular cost pillar breakdown allows policymakers to identify investment priorities. Several limitations must be acknowledged. First, the model assumes that importation scales with DRC prevalence using a fixed scaling factor (56%), whereas real importation risk likely varies with border activities, seasonal factors, and individual mobility patterns. A forthcoming metapopulation model incorporating movement data and border crossing statistics will address this limitation. Second, the cost per case for the rapid response was derived from a scenario with 26–31 cases; if the rapid response occasionally fails, the cost per case would drop, making the rapid response even more cost-saving. Third, we did not include indirect economic costs such as lost trade or event cancellations; including those would further favour the rapid response. Fourth, the model does not account for spatial heterogeneity. Fifth, the Sobol analysis used 300 parameter sets due to computational constraints; indices near zero had wide confidence intervals and some first-order indices were negative (sampling artefacts). Sixth, the behavioural response is modelled as a function of daily reported incidence only; in reality, behaviour also changes in response to media and trust [34]. Finally, the HCW infection probability (0.4286) was derived from a small initial cluster; larger datasets would refine this estimate. Future work should focus on component-wise decomposition of the 7-1-7 framework to determine which element (detection acceleration, contact tracing, quarantine, behavioural response) delivers the largest marginal benefit. Age-structured models with school contact matrices are needed to assess EVD spread in educational settings. A full cost-benefit analysis of cancelling mass gatherings (e.g., Martyrs Day) would help policymakers balance public health and economic objectives. Finally, the forthcoming metapopulation model will provide real-time importation risk estimates to optimise screening strategies at the eight high-risk ports of entry identified by IOM.

## 5 Conclusion

A rapid outbreak response aligned with the 7-1-7 framework (using Uganda’s achieved 3-1-5 timeline) is projected to reduce cases by 60–66% and deaths by 62–63% compared to a delayed counterfactual, while being strictly dominant (cost-saving and life-saving) under importation risks up to 56%. The case fatality rate, import probability, and basic reproduction number are the dominant drivers of uncertainty, but detection delay retains important interaction effects. Institutionalising the 7-1-7 framework in Uganda with sustainable domestic financing, community-based surveillance at unofficial border points (especially the eight high-risk ports of entry), three-consecutive-PCR laboratory capacity, and multilingual risk communication is a cost-saving investment that should be prioritised. Regional synchronisation of 7-1-7 targets across East Africa would further enhance collective health security.

## Contributors

AWW, LN, SPS, PM, MB, WS, AK, BL, MN, PAI, PK, CO, ANM, MMM and DA conceived the study and developed the model. AWW performed the analyses and DALY calculations. SPS and PM reviewed and revised model assumptions and parameterisation. PM, MB, WS, AK, BL, MN, PAI, PK, CO, ANM, DA and MMM provided government policy insights and first-hand data. All authors interpreted the results and approved the final manuscript.

## Data availability

All aggregated simulation outputs (median cases, deaths, costs, DALYs) are presented in Table 2. No primary data were collected; all input parameters were obtained from published literature and public reports cited in the manuscript.

## Code availability

The R code for the model and Sobol analysis is available at https://github.com/abelwalekhwa/bundibugyo-717-model.

